# Copy number signatures in cervical samples enable early detection of high-grade serous ovarian carcinoma

**DOI:** 10.1101/2025.04.23.25325778

**Authors:** Laura Martin de la Fuente, Srinivas Veerla, Minerva X. Li, Guyuan Tang, Anna Ebbesson, Laura Mannarino, Lara Paracchini, Sergio Marchini, Maurizio D’Incalci, Anna Måsbäck, Susanne Malander, Päivi Kannisto, Ingrid Hedenfalk

## Abstract

**Background:** Ovarian cancer is often diagnosed in advanced stages, resulting in poor outcomes. There is an unmet need for a sensitive and specific screening tool for early-stage detection of ovarian cancer.

**Methods:** Recognizing that high-grade serous ovarian carcinoma (HGSC) is driven by copy number alterations (CNAs) and that tumor DNA can be detected in cervical samples, we analyzed CNAs from shallow whole genome sequencing of 212 cervical samples from 128 women with/without HGSC, including 29 germline *BRCA1/2* mutation carriers. Using a machine-learning classifier, we developed **H**igh-grade serous ovarian cancer **C**ervical copy number **sig**nature (HCsig), a predictor for HGSC detection.

**Results:** HGSC-derived CNAs were detectable with HCsig in cervical samples collected several years before diagnosis. Importantly, HCsig correctly identified HGSC in 79% of archival cervical samples, including 91% stage I-II (0-27 months pre-diagnosis), and 77% stage III-IV (0-65 months pre-diagnosis). Among patients with HGSC who had multiple pre-diagnostic samples collected during the pre-symptomatic phase, 85% had at least one HCpositive cervical sample before surgery (up to 65 months before diagnosis). Detection rates were 90% and 76% for *BRCA1/2*-mutated and wildtype HGSC, respectively. Validation in 172 independent samples (0-98 months pre-diagnosis) showed 76% sensitivity and 94% specificity (AUC=0.83), including high sensitivity for early-stage cancers.

**Conclusions:** We show that applying the HCsig classifier to cervical samples, including from non-symptomatic women several years before diagnosis, holds promise for early-stage detection and secondary prevention of HGSC. Moreover, it may serve as a screening tool to aid decision-making regarding timing of risk-reducing surgery in high-risk populations.

## BACKGROUND

Almost two-thirds of women diagnosed with ovarian cancer die from their disease (1). One of the main reasons for the high mortality rate is the lack of effective screening methods leading to diagnosis in advanced stages. As women detected with localized disease have a survival rate >90%, considerable efforts are being invested in developing effective screening tools for early detection.

The majority of high-grade serous tubo-ovarian carcinomas (HGSC) are derived from precursor lesions arising from epithelial cells in the fimbriated end of the fallopian tubes (2). Pre-cancerous lesions, so-called serous tubal intraepithelial carcinomas (STICs) harbor clonal *TP53* mutations and display a histological appearance resembling HGSC (3) (4). Dysplastic cells harboring mutant *TP53* (p53 signatures) are believed to precede the STIC lesions (5) (6). These findings provide evidence for the possibility of finding tumor-driving mutations in asymptomatic women with early-stage HGSC (7). To this end, Kinde *et al.* showed that somatic mutations in DNA shed from ovarian cancers could be detected in standard liquid-based Pap test specimens collected at the time of diagnosis by massively parallel sequencing (8). More recently, we and others have reported the detection of tumor-specific *TP53* mutations in archival liquid-based cervical cytology samples obtained from asymptomatic women up to several years prior to the HGSC diagnosis using ultra-sensitive methods (9) (10). However, it has recently been shown that very low-frequency, cancer-like, age-related *TP53* mutations are found in healthy women, revealing the challenge of using mutations in *TP53* for the early detection of ovarian cancer (11) (12). Hence, while sensitivity has been the main constraint in previous studies, specificity has emerged as the major challenge for early detection using ultra-sensitive sequencing approaches.

HGSC is a prototypical tumor characterized by genomic instability, where mutational processes acting on cancer genomes drive copy number alterations (CNAs) that can be readily investigated by whole genome sequencing (WGS) (13). In fact, evolutionary analyses have reported that aneuploidy and genomic instability, in addition to *TP53* mutations, can occur as early in HGSC progression as in the STIC lesions (14). To address the possibility of detecting the earliest steps in the evolution of HGSC using an unbiased approach, we therefore applied low-pass/shallow WGS (sWGS) to assess the consequence of the earliest known molecular event in the development of HGSC, mutations in *TP53*, leading to genomic instability. In this study, we interrogated genome-wide CNAs in a discovery cohort of 309 samples, including 212 cervical samples collected at surgery and archival samples from routine screening for cervical cancer taken up to 94 months prior to surgery from HGSC patients, *BRCA1/2* mutation carriers undergoing risk-reducing salpingo-oophorectomy (RRSO) and women with benign gynecological conditions. We explored genomic landscapes in the cervical samples using copy number signatures derived from HGSC (15) as well as pan-cancer copy number signatures for chromosomal instability (16) (17). To derive novel copy number signatures more fitting for low DNA-abundance cervical samples we used the unsupervised, machine learning-based clustering method Systematic Random forest Integration with Qualitative threshold (SRIQ) (18) to develop the **H**igh-grade serous ovarian cancer **C**ervical copy number **sig**nature (HCsig) classifier. A prediction model was constructed based on centroids for HCsig, which was tested and validated in an independent cohort of 172 samples from HGSC and healthy women (**Figure 1**). Our findings demonstrate the feasibility of applying sWGS to standard cervical samples for simple, minimally invasive, and accurate early detection of HGSC, with the potential for downstaging and improved survival. As a screening tool for risk assessment in high-risk populations, *e.g.* women with germline mutations in *BRCA1/2*, HCsig may support clinical decision-making regarding the timing of RRSO.

**Figure 1.**
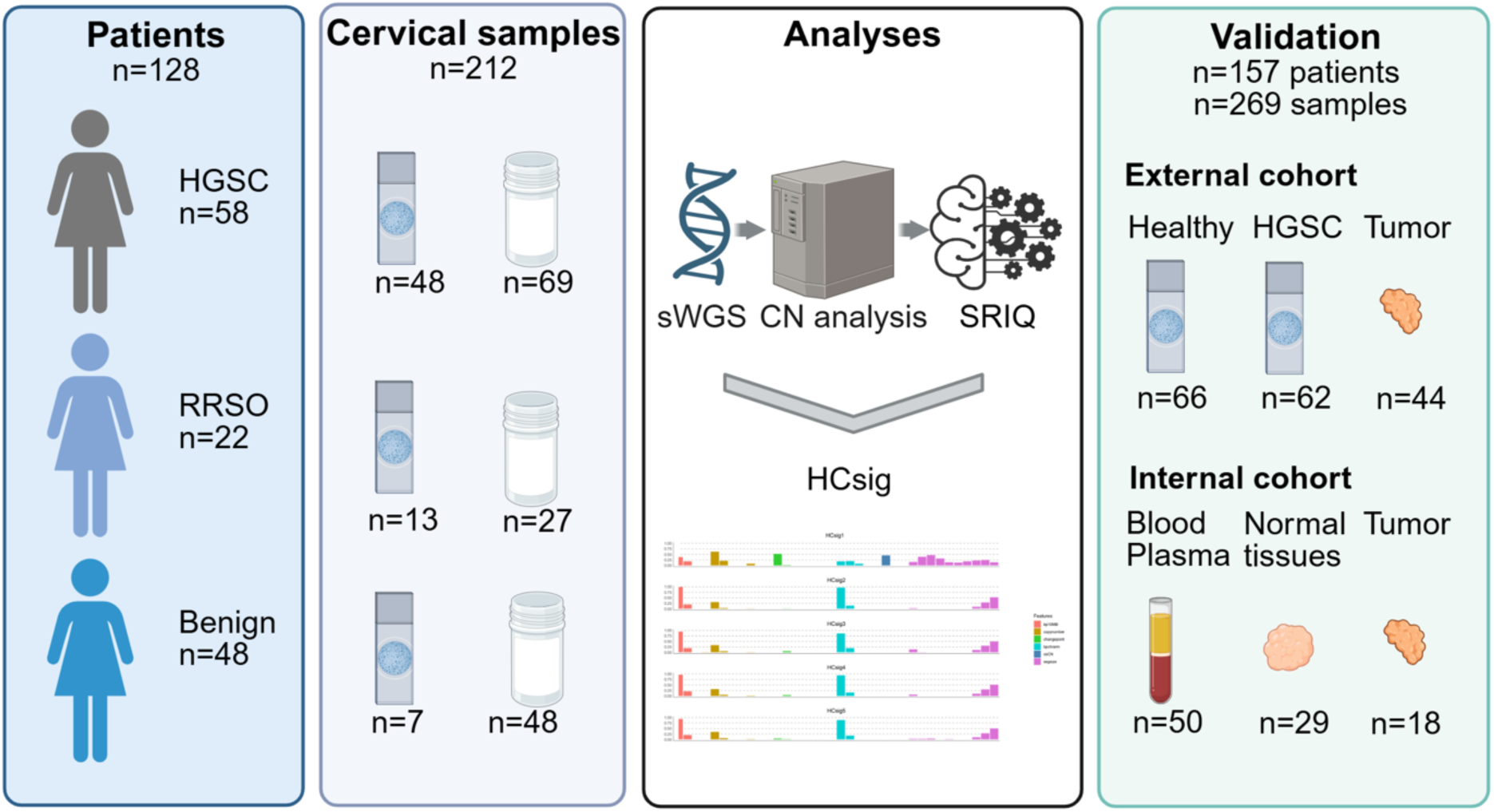
Overview of patients, samples and workflow for HCsig. Liquid- and slide-based cervical samples from women with HGSC, *BRCA1* and *BRCA2* mutation carriers undergoing risk-reducing salpingo-oophorectomy (RRSO) and women with benign gynecological conditions were used for sWGS. Cervical samples were from 0-94 months before surgery. The random forest machine learning-based clustering method SRIQ was applied to define HGSC-derived copy number signatures, HCsig 1-5. Validation was performed using an independent cohort of women with HGSC and healthy controls (external cohort), as well as normal tissues, tumors and blood/plasma from the discovery cohort (internal cohort). Created using BioRender.com.

## METHODS

### Study design

The discovery cohort in the present study included 128 patients and 309 samples: 173 samples from 58 women diagnosed with HGSC, 82 samples from 48 women diagnosed with benign gynecological conditions and 54 samples from 22 *BRCA1/2* mutation carriers who underwent RRSO. Clinical information and sample types by patient group are presented in **Table 1**.

**Table 1.** Patient and sample information.

|  | HGSC | RRSO | Benign | Total |
| --- | --- | --- | --- | --- |
| **Patients (n)** | **58** | **22** | **48** | **128** |
| Age Range (years) | 43-77 | 36-73 | 23-86 |  |
| Median (years) | 64 | 45 | 66 |  |
| *BRCA1/2* Mutant (n) | 10* | 22 | 0 | **32** |
| Wildtype (n) | 42 | 0 | 0 | **42** |
| Not tested (n) | 6 | 0 | 48 | **54** |
| Stage Advanced (FIGO III and IV) (n) | 47 | N/A | N/A |  |
| Localized (FIGO I and II) (n) | 11 | N/A | N/A |  |
| CA-125 Range (U/mL) | 7.4-11,666<sup>†</sup> | <sup>†</sup> | 4.1-1,207<sup>†</sup> |  |
| Median (U/mL) | 925 | N/A | 35 |  |
| **Samples (n)** | **173** | **54** | **82** | **309** |
| **Cervical samples (n)** | **117** | **40** | **55** | **212** |
| Diagnostic (n) | 31 | 17 | 55 | 103 |
| Archival (n) | 86 | 23 | 0 | 109 |
| **Additional samples (n)** | **56** | **14** | **27** | **97** |
| Tumor tissue (n) | 18 | N/A | N/A | 18 |
| Normal tissue<sup>‡</sup> (n) | 9 | 7 | 13 | 29 |
| Blood (n) | 18 | 4 | 7 | 29 |
| Plasma (n) | 11 | 3 | 7 | 21 |

HGSC, high-grade serous ovarian cancer; RRSO, risk-reducing salpingo-oophorectomy
\*, n=6 germline *BRCA1* mutations, 1 germline *BRCA2* mutation, 3 somatic *BRCA1* mutations
<sup>†</sup>, missing CA-125 values: n=2 HGSC, 22 RRSO, 12 benign
<sup>‡</sup>, endometrium, ovary, uterus
N/A, not applicable

Two in-house cohorts contributed to the discovery cohort; one prospective cohort where diagnostic cervical samples were collected from 187 women who underwent RRSO or had surgery due to adnexal masses at the Gynecological Department in Lund, Skåne University Hospital, Sweden (2015–2017) (9) and one cohort where 59 patients with HGSC or RRSO surgery (2017–2020), with accessible archival cervical samples, were retrospectively identified. To be eligible for this study, the availability of archival samples was also required from the patients in the first cohort. A flowchart of the inclusion and exclusion of patients is shown in **Figure S1**. The study was approved by the Ethics Committee at Lund University, Sweden.

### Cervical samples

Archival cervical samples from cervical cancer screening were obtained from the Pathology Department at Skåne University Hospital, Sweden, while fresh-frozen tissue, blood, and plasma samples were available from the regional biobank for gynecological conditions in the southern Swedish healthcare region. Liquid-based cervical cytology samples were preserved in either DNAgard® (Sigma-Aldrich), ThinPrep® solution (Hologic) or SurePath® solution (BD Biosciences). Diagnostic samples were collected at surgery, and archival samples were from any time before surgery (range: 0 to 94 months).

In total, we analyzed 117 cervical samples from the HGSC group: 31 diagnostic cervical samples (from 19 patients), and 86 archival samples (from 52 patients). For the RRSO group, we analyzed 40 cervical samples: 17 samples were collected at surgery (from 12 patients), and 23 were archival (from 14 patients). Fifty-five diagnostic cervical samples were from women with benign gynecological conditions (48 women). Twenty-five patients (20 HGSC and five RRSO) had samples available from multiple time points. In addition, blood, plasma, tumor and normal tissues were also collected, for a total of 309 samples from 128 patients (**Table S1**).

### Sample processing and library preparation

The cover glass or film was removed from cervical cytology slides using xylene or acetone, respectively, and DNA was extracted with the Maxwell RSC DNA FFPE kit (Promega). Liquid-based cervical samples were extracted with the QIAamp DNA Micro kit (Qiagen). For the additional samples, DNA was extracted using the AllPrep DNA/RNA Mini kit (fresh-frozen tissue) or QIAmp DNA Blood Midi kit (blood and plasma).

DNA concentrations were measured by Qubit fluorometric assays and quality-checked by Illumina TruSeq FFPE Library Prep QC Kit (Illumina). The QIAseq FX DNA library prep kit (Qiagen) was used for library preparation according to the manufacturer’s protocol. Briefly, sequencing libraries were prepared from 1-10 ng DNA with 4-12 cycles of amplification depending on the DNA input. qPCR with EvaGreen DNA-binding dye was used to monitor library enrichment. Low pass/shallow whole genome sequencing (sWGS) was performed on the NovaSeq 6000 platform (Illumina) at the Center for Translational Genomics (CTG) in Lund, Sweden. The target sequencing depth was >1X for all sample types, except blood and plasma, which were sequenced at 0.5X. Approximately 15% of the extracted cervical samples were not sequenced due to poor DNA quality and/or quantity.

### Data analysis workflow

A data analysis workflow was assembled to extract copy number features from all samples. We calculated similarities with previously published cancer-derived copy number signatures and generated novel copy number signatures from HGSC in cervical samples using sWGS data. Demultiplexed FASTQ files from paired-end sWGS data were used as input. The workflow contains copy number analysis to generate relative copy number profiles and solutions for cellularity and ploidy, as well as the manually curated segmentation bin sizes for each sample. Absolute copy number profiles were generated and used to explore HGSC-derived copy number signatures (CNsignatures) (15), pan-cancer chromosomal instability copy number signatures (CIN signatures) (16), and allele-specific copy number signatures (panConusig) (17). For the construction of the HGSC-specific cervical copy number prediction model HCsig, we used the absolute copy number profiles of the cervical samples from patients with and without HGSC, performed unsupervised SRIQ clustering (18) to generate copy number-based signatures and subsequently a prediction model, which was then tested and validated using an independent dataset from Paracchini *et al.* (19).

### Absolute copy number analysis

Raw sequencing data were demultiplexed using Illumina’s bcl2fastq software to generate FASTQ files, which were subsequently aligned to the reference genome (hg38) using BWA (v0.7.17-r1188). The alignment files were processed with Samtools (v1.13) to convert SAM to BAM format, followed by duplicate marking using PicardTools (v3.3.0), and GATK software (v4.1) was applied for base quality score recalibration (BQSR).

Relative copy number profiles were generated using a modified version of QDNAseq (20). Custom bin annotations for the hg38 reference genome were created to optimize the analysis for a 100bp paired-end sequencing setup, following the package’s recommended workflow. A new control dataset, comprising 58 human genomes from the 1000 Genomes Project Phase 3, was incorporated to enhance analytical accuracy.

The segmented relative copy number profiles, derived under manually optimized bin sizes for the 309 samples, were processed using the Rascal R package. Rascal employs the ACE (Absolute Copy-number Estimation) method to scale relative copy numbers to absolute values. To refine the results, we applied an additional model adjustment using mutant allele frequencies (MAFs) for *TP53*, previously quantified via ddPCR (IBSAFE®, Saga Diagnostics) in three patients (9).

### Integration with published copy number signatures

The outputs generated by Rascal were utilized to derive sample-by-component matrices for HGSC-specific copy number signatures (15) and pan-cancer chromosomal instability (CIN) signatures (16). We performed a cosine similarity analysis to identify signatures with the most similar patterns in our dataset. This compared the sample-by-component matrices with the corresponding signature-by-component definition matrices, resulting in sample-by-signature matrices containing cosine similarity values.

For cases with matched blood samples, we applied panConusig (17), which leverages allele-specific copy number profiles. Prior to calculating these profiles, we used a modified workflow of Battenberg v2.2.9 on the BAM files to generate haplotype-phased data. The workflow was adapted to include only the initial steps: counting the major and minor allele frequencies and determining phased states. The haplotype-phased allele frequency data were subsequently processed with the shallow-coverage version of ASCAT (ASCAT.sc v0.1) to calculate allele-specific copy number profiles.

### Construction of HGSC-derived copy number signatures (HCsig) using cervical samples

Absolute copy number profiles in cervical samples from women with HGSC were used to extract genome-wide copy number features. These features included segment sizes (segsize), breakpoint counts per 10Mb (bp10MB), lengths of oscillating copy number segment chains (osCN), copy number change points (changepoint), segment copy numbers (copynumber), and breakpoint counts per chromosome arm (bpchrarm). Each feature was further decomposed into distinct components using mixture modeling, as described in (15), resulting in 36 unique components. These components formed the basis of a sample-by-component matrix. This matrix was then used as input for the SRIQ algorithm (18), an unsupervised clustering approach that integrates principles from the Random Forest machine learning classifier, quality thresholding, and k-nearest neighbor clustering. The aim of the SRIQ method is to delineate a core cluster of samples or components sharing common patterns without the need for *a priori* knowledge of the data or a predetermined number of clusters (K). As a result of this analysis, distinct clusters were successfully identified. For each resulting cluster, a centroid was computed. The centroids represent the mean vector (or average components profile) of all samples within the cluster, effectively capturing the dominant alteration pattern or signature characteristic of that cluster and were used to create the HCsig classifier.

### Validation of the centroid-based prediction tool HCsig in independent data

The HCsig signature profiles were used to develop and validate a prediction model for identifying HGSC in an independent cohort comprising 172 samples. This cohort included 44 biopsy samples, and 128 cervical samples collected from presymptomatic women, with 62 samples from individuals diagnosed with HGSC and 66 from healthy controls (19). The downloaded BAM files, which were processed using genome reference hg38, were run through our analysis pipeline to produce a sample-by-component matrix. Each sample was compared to the HCsig centroids using Spearman correlation. Samples were then annotated with the HCsig signature showing the highest Spearman correlation value. Samples with close similarity to two centroids, one of which being HCsig1 (delta<0.05 between HCsig1 and any other HCsig) were considered as HCpositive. A custom Java-based tool (see Code Availability section) was developed for this analysis. The model’s performance was evaluated using a receiver operating characteristic (ROC) curve, with the area under the curve (AUC) providing an overall measure of predictive accuracy. Additionally, confusion matrix metrics, including sensitivity, specificity, and accuracy, were calculated to assess the model’s ability to classify samples accurately as originating from HGSC patients or healthy controls. All performance analyses, including calculations and visualizations, were conducted using Python’s scikit-learn package.

### Fraction of Genome Altered (FGA) analysis

To calculate FGA, we used segmented data derived from the sWGS analysis. Genomic segments were classified as altered if their log2 ratio exceeded an absolute threshold (set at ±0.2). Next, we summed the genomic length of all altered segments. Finally, FGA was computed by dividing the total length of altered regions by the total autosomal genome size (excluding sex chromosomes). For classification purposes, samples with an FGA of 20% or higher were classified as positive.

### Statistical analyses

Chi-square tests were used for categorical variables. P<0.05 was considered statistically significant. Confidence intervals (95% CI) were calculated using the Normal Approximation method. Cosine similarity was used to quantify the closeness between different signature exposure profiles. Spearman correlation was used to assign HCsig signatures to samples in the validation cohort.

## RESULTS

### sWGS and cancer-derived copy number signatures in cervical samples, tumors and normal tissues

The discovery cohort consisted of cervical samples (slide- and liquid-based, collected at diagnosis, and up to 8 years before diagnosis), tumors, blood/plasma and normal tissues from a previously published prospective cohort (9), and a retrospective cohort (total n=58 HGSC, 22 RRSO and 48 with benign gynecological conditions). The patient cohorts and sample types are shown in **Table 1**. Information on *BRCA1* and *BRCA2* (henceforth *BRCA1/2*) mutation status, FIGO stage and CA-125 levels was retrieved from patient records. The majority of the HGSC patients were diagnosed with advanced-stage disease (47/58, 81%). We applied sWGS to all samples, aiming at a coverage of approximately 1X (range 0.4-2.2X). Although the DNA quality from slides was poorer than the liquid-based cervical samples, the results of the library preparation and sequencing were satisfactory. The average number of mapped reads was 49.2x10^6^, 25.4x10^6^, 19.8x10^6^, 41.7x10^6^, 40.6x10^6^, 57.9x10^6^ for tumor, blood, plasma, normal tissues, slide-based, and liquid-based cervical samples, respectively (range: 27.2x10^6^-63.4x10^6^, 18.1x10^6^-41.8x10^6^, 16.5x10^6^-24.8x10^6^, 32.1x10^6^-64.1x10^6^, 12.7x10^6^-55.3x10^6^, 34.0x10^6^-105.0x10^6^) (**Figure S2**). Various fixed bin sizes were explored based on sequencing quality using QDNASeq (20), with a final selection of 50kb for all sample types. The generation of relative and absolute copy numbers, and the assessment of copy number signatures, was performed as outlined in the Methods section.

Signature similarity exposure profiles from three previously published collections of cancer-derived copy number signatures (15) (16) (17) were evaluated to identify copy number patterns across sample types, including cervical samples (diagnostic samples from the time of surgery and archival samples collected during routine screening for cervical cancer), tumor and normal tissue, blood and plasma from women with HGSC, women at high risk of developing HGSC due to germline *BRCA1/2* mutations and women with benign gynecological conditions (**Table S2**). The HGSC-derived CN signatures (15) more readily captured CNAs in cervical samples from HGSC patients in our cohort compared to the pan-cancer signatures (**Figure S3, Table S1**). Overall, HGSC and pan-cancer signatures defined by a high number of breakpoints, short to medium-sized, clustered and oscillating copy number changes, proposed to be caused by impaired homologous recombination, replication stress and/or chromothripsis, were commonly observed in tumors and cervical samples from patients with HGSC. In addition, the allele-specific panConusig signatures also captured LOH events in both tumors and a subset of cervical samples.

### Collation and exploration of HGSC-derived copy number signatures using cervical samples

The previously reported copy number signatures were derived from advanced-stage cancers (15); therefore, we hypothesized that they may lack the sensitivity required for capturing the early stages of HGSC in cervical samples. With the aim of developing a screening test for the early detection of HGSC using cervical samples, we decided to use the cervical samples in our discovery cohort to generate copy number signatures indicative of HGSC based on the six fundamental copy number features previously reported as hallmarks of genomic aberrations (15), and found in both HGSC tumor tissue and cervical samples in our cohort.

Cervical samples from the HGSC group (n=117) and the benign and RRSO groups (n=55 and 40, respectively) were included in the analysis. To generate HCsig, a copy number-based classifier for HGSC using cervical samples, we applied the unsupervised, random forest machine learning-based clustering method SRIQ (18) to discover clusters of samples in our data and to identify the copy number features defining these clusters. SRIQ identified five discrete copy number clusters (signatures), with the majority of the HGSC cervical samples being enriched for HCsig1. The heatmap in **Figure 2A** displays the five clusters and their defining features in each sample, and the bar plot in **Figure 2B** shows the weights of the defining components in the signatures, denoted HCsig 1-5.

**Figure 2.**
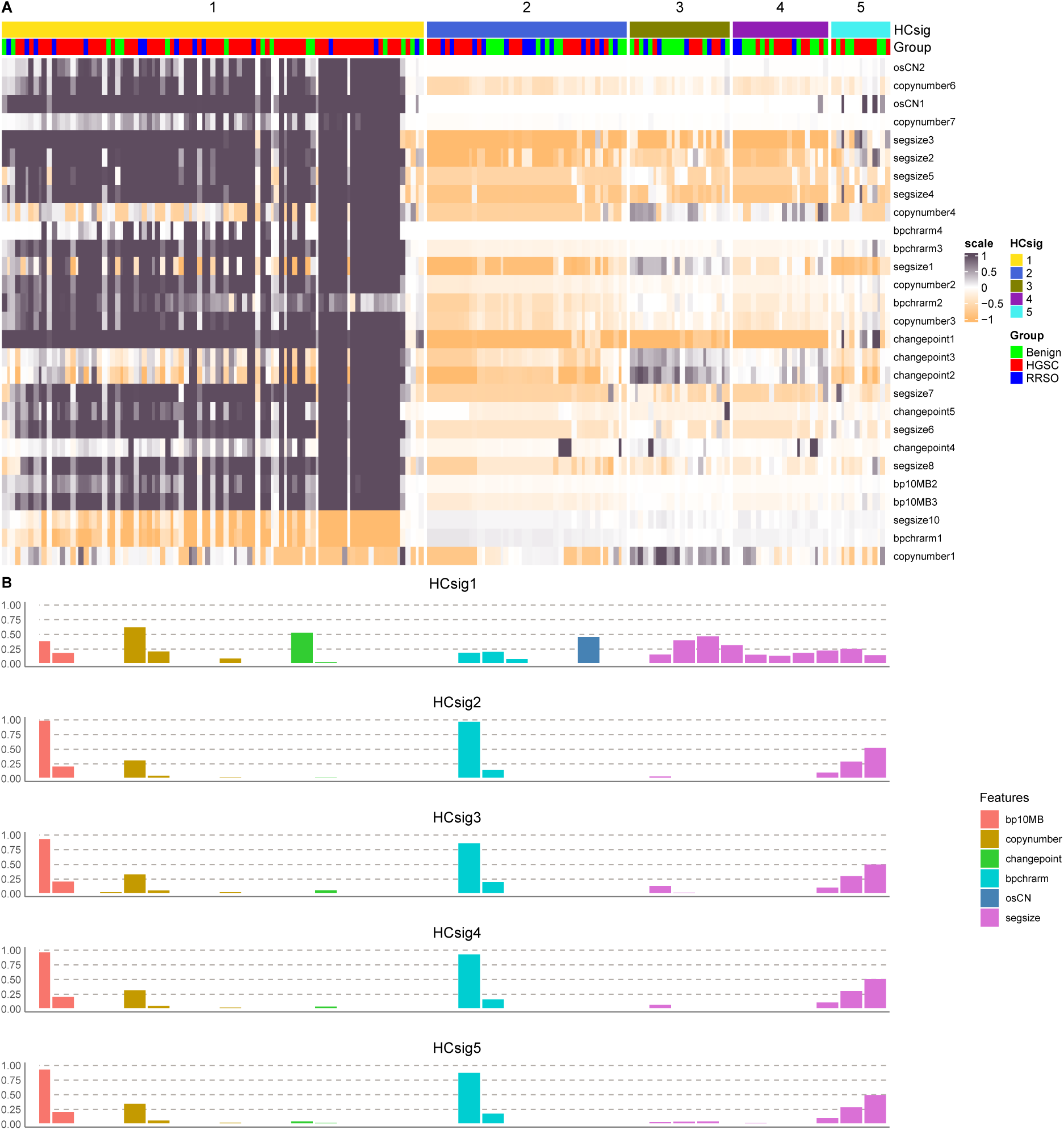
Copy number features underlying the HCsig classifier in cervical samples. **(A)**. SRIQ-based clustering and heatmap of cervical samples from patients with HGSC (n=117), benign gynecological disease (n=55) and who had RRSO (n=40). Samples are clustered on the x-axis, with the HCsig clusters 1-5 and patient groups shown above. The copy number features are shown on the y-axis. (**B).** Bar plot displaying the distributions and weights of the defining copy number components for HCsig 1-5.

The majority (67/117, 57%) of the HGSC cervical samples clustered together and exhibited enrichment for HCsig1, defined by highly variable segment sizes, oscillating copy number events, one to two breakpoints per chromosome arm, breaks evenly distributed across the genome, single copy number change points and copy number two. Notably, HCsig1 shared many features with the HGSG-derived CN signatures s3, s6 and s7 related to homologous recombination deficiency (HRD) and failure of cell cycle control (15), and was identified as a descriptor for HGSC in the HGSC cervical samples in our cohort.

HCsig 2-5 demonstrated few defining copy number features and limited differences; these signatures were mainly defined by large segment sizes and few breakpoints. The majority of the cervical samples from women with benign conditions and from women who had undergone RRSO were enriched for HCsig 2-5, suggesting that large segment sizes and the occurrence of few breakpoints are not tumor-specific features. Taken together, samples enriched for HCsig1 (*i.e.* if HCsig1 was the most highly enriched signature, or, alternatively, if HCsig1 was the second-most enriched signature with a delta <0.05 compared to the highest signature, see Methods) were considered as HCpositive.

We next examined whether our sWGS approach could be used to capture HGSC in archival cervical samples, mirroring the pre-diagnostic/pre-symptomatic setting. The HCsig clusters were converted into centroids and used to perform a binary centroid-based prediction analysis: HCpositive or HCnegative. In total, 46/58 (79%) women with HGSC had at least one positive cervical sample before surgery. Liquid and slide-based cervical samples showed similar distributions of HCsig 1-5 and similar fractions of HCpositive and HCnegative samples, indicating limited impact of sample processing (**Figure S4**). Importantly, 91% (10/11) of women with stage I-II HGSC had at least one positive cervical sample before surgery (range: 0-27 months), while 77% (36/47) of women with advanced stage disease (stage III-IV) had at least one positive cervical sample before surgery (range: 0-65 months). Notably, for women with *BRCA1/2*-mutated tumors (germline or somatic), 90% (9/10) were detectable in at least one pre-diagnostic cervical sample (range: 0-54 months), compared to 76% (32/42) of the *BRCA1/2* wildtype tumors (range: 0-51 months). Among healthy women in the benign group, 9/48 (19%) had a false positive cervical sample, while 11/22 (50%) *BRCA1/2* germline mutation carriers who had an RRSO, one of whom displayed STICs according to the pathology reports, had an HCsig positive sample (**Figure 3**). There were no differences in exposure to HCsig 1-5 between cervical samples from *BRCA1/2*-mutated HGSC and those from women with sporadic HGSC (**Figure S5**). To further explore the distribution and origin of the features identified in the HCsig signatures, we investigated the exposures across different sample types using cosine similarity, clearly demonstrating enrichment of the HCpositive class in the majority of tumor tissue samples (15/18, 83%), validating the HGSC origin of HCsig1. Only 3/18 (16%) blood and 4/11 (36%) plasma samples from HGSC patients were HCpositive, indicating lower sensitivity in peripheral samples compared to cervical samples (**Figure 3**).

**Figure 3.**
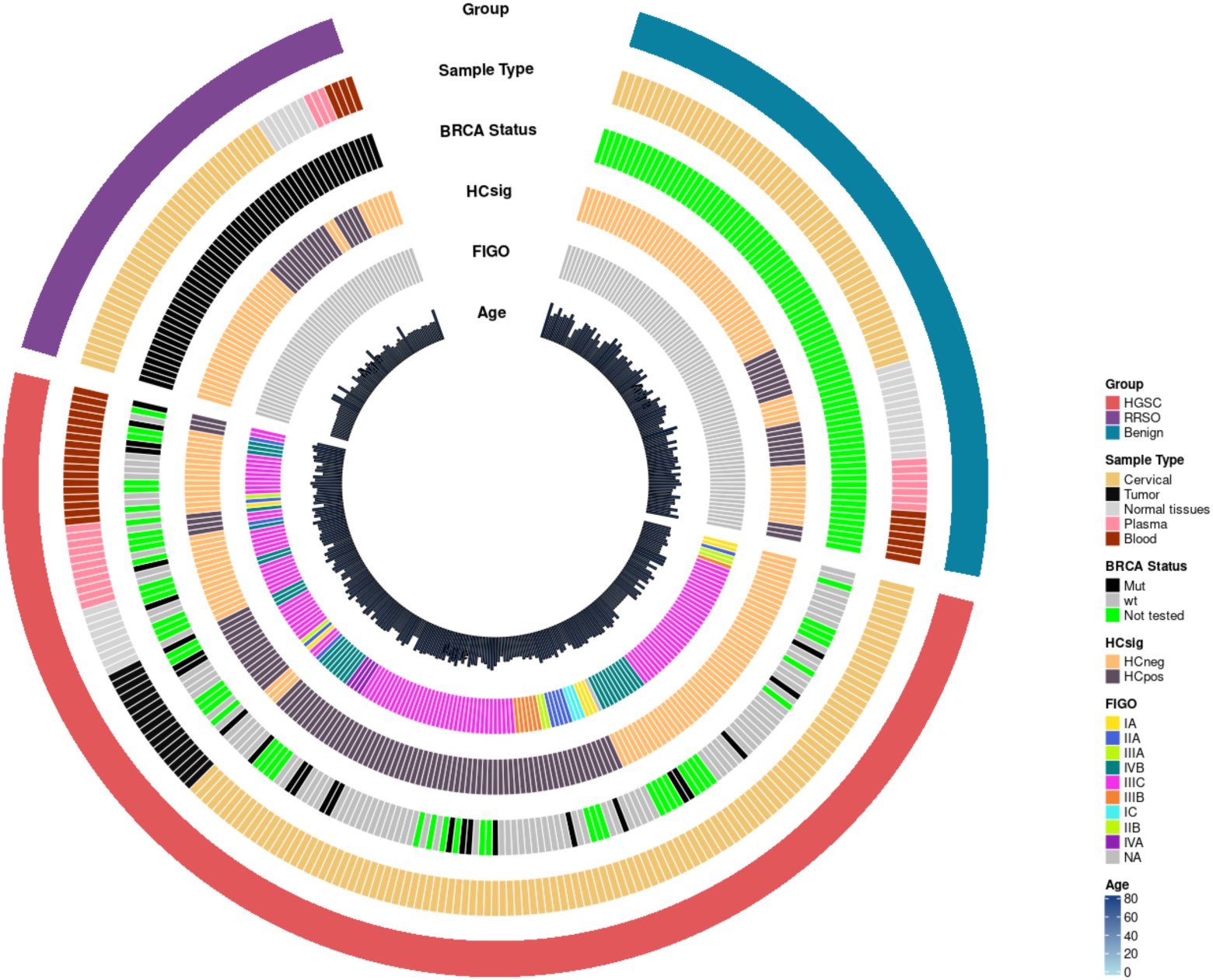
Circular annotation plot summarizing clinical, pathological and molecular features and HCsig across all samples (n = 309), including cervical (68.6%) and non-cervical (31.4%) samples. The outer ring represents the disease group, showing the distribution of HGSC (56.0%), RRSO (17.5%), and benign (26.5%) cases. The next ring indicates sample type, including cervical (68.6%), tumor (5.8%), normal tissues (9.4%), plasma (6.8%), and blood (9.4%) samples. *BRCA1/2* mutation status is represented in the following ring, with mutated (25.9%), wild-type (31.4%), and not tested (42.7%) cases. The fourth ring depicts HCsig, derived from our genomic classifier, showing HCpositive (41.1%) and HCnegative (58.9%) cases. The fifth ring encodes FIGO stage at HGSC diagnosis spanning early to advanced disease: IA (2.3%), IC (1.0%), IIA (3.6%), IIB (0.3%), IIIA (1.6%), IIIB (2.3%), IIIC (34.0%), IVA (1.3%), IVB (9.4%), and NA (44.3%). Note: Excluding samples from the benign and RRSO groups, the distribution is: IA (4.0%), IC (1.7%), IIA (6.4%), IIB (0.6%), IIIA (2.9%), IIIB (4.0%), IIIC (60.7%), IVA (2.3%), IVB (16.8%), NA (0.6%). The innermost ring displays patient age using a bar plot and color gradient from light blue to dark blue, with age groups distributed as follows: 20–30 (0.3%), 30–40 (1.3%), 40–50 (23.6%), 50–60 (15.9%), 60–70 (33.0%), 70–80 (24.6%), and 80–90 (1.3%) years, respectively.

When including all sample types from all patient groups in the analysis, the sensitivity of the HCsig classifier was 86%, the specificity 79% and the accuracy 82%. The normal samples (blood, plasma and normal tissues) displayed similar exposures to all HCsig signatures, *i.e.* no enrichment of tumor-like features. There were no indications in the pathology reports specifying the presence of invasive features in tissues from women with an HCpositive sample from the benign or RRSO groups. However, the fallopian tube tissue from one of the women who had undergone an RRSO was found to display a STIC lesion. Taken together, the most distinguishing features captured in HCsig1 (HCpositive) in cervical samples from HGSC patients were also more prevalent in tumor tissue compared to normal tissues.

### Modelling pre-symptomatic detection: analysis of HCsig in longitudinal cervical samples from HGSC patients

To explore the ability of the HCsig classifier to capture copy number features from HGSC in cervical samples collected during routine gynecological examination of pre-symptomatic women and who were later diagnosed with HGSC, we applied the HCsig classifier to archival cervical samples to simulate the pre-symptomatic setting. Women later diagnosed with HGSC, who had ≥2 cervical samples taken at different timepoints were included in the analysis (n=20 patients) and samples were ordered according to the time of sampling in relation to the time of diagnosis (range: 0-94 months).

The HCsig classification of cervical samples from women diagnosed with HGSC sorted by time is shown in **Figure 4**. Enrichment for the HCpositive class was observed in both diagnostic and archival cervical samples from women with HGSC. Seventeen of 20 patients with HGSC (85%) had at least one HCpositive cervical sample before surgery (up to 65 months before diagnosis).

**Figure 4.**
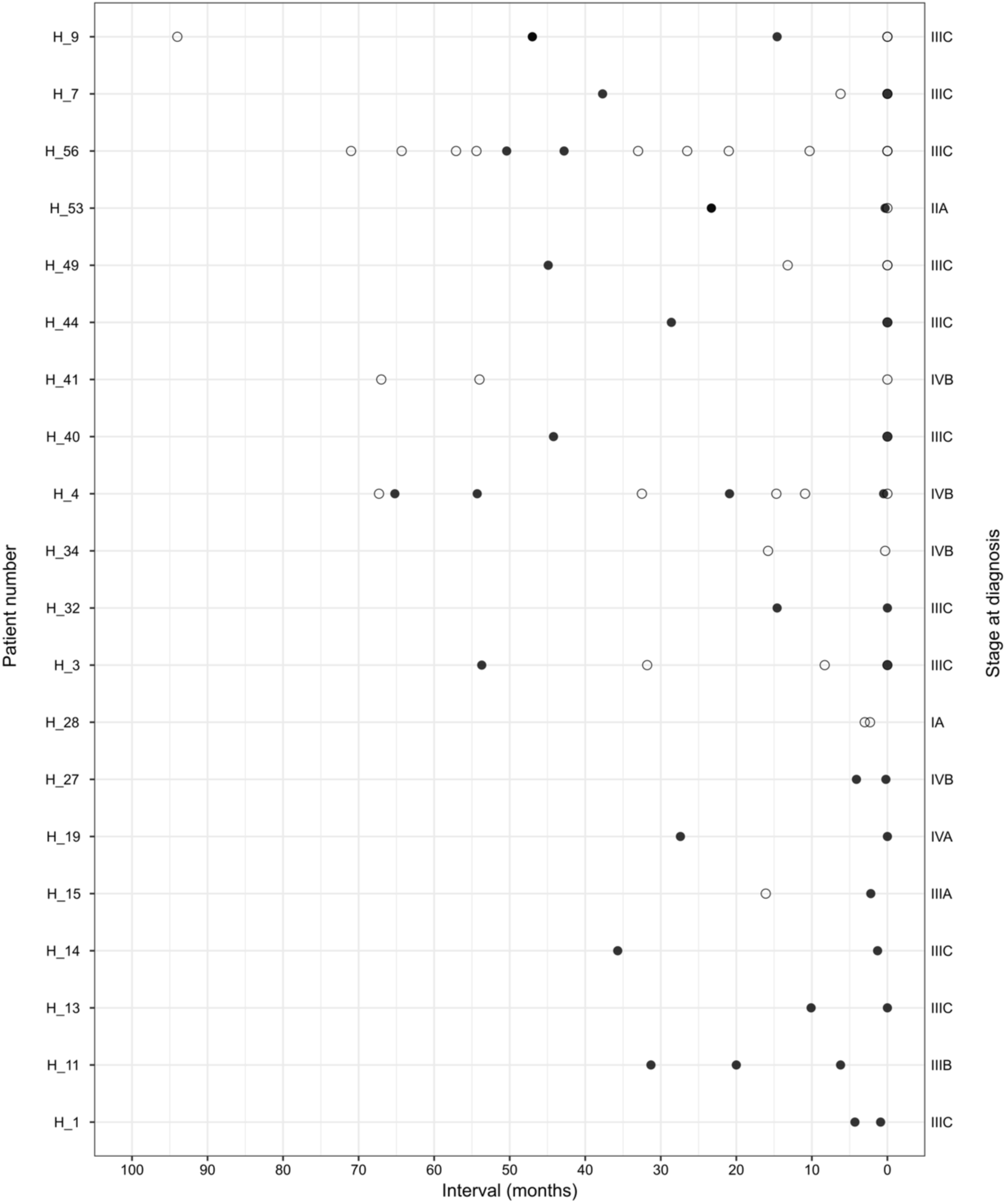
HCsig classification in longitudinal cervical samples from women with HGSC. The dot plot shows the classification of cervical samples from women with HGSC and ≥2 cervical samples available. The x-axis shows the time in months before diagnosis (right to left). Filled circles represent HCpositive samples and open circles represent HCnegative samples. FIGO stage at diagnosis of HGSC is indicated for each patient on the right y-axis.

Enrichment for the HCpositive class in archival samples obtained up to >5 years before diagnosis supports the notion that copy number features captured from sWGS data may enable early detection of HGSC in pre-symptomatic women. In summary, these findings show that complex CNAs resembling those observed in HGSC and captured using HCsig can be detected in archival cervical samples obtained from asymptomatic women up to several months, and even years, prior to the onset of symptoms and subsequent diagnosis.

### Validation of the HCsig classification model for HGSC using samples from an independent dataset

Finally, to examine the diagnostic potential of our classification model, we performed a validation using an independent dataset with a comparable number of samples and patients, including archival cervical samples from women with HGSC and healthy women. The time from the collection of cervical samples to the time of HGSC diagnosis ranged from 0 months to 13.6 years (19). sWGS data (1-5X) from 172 samples were preprocessed and analyzed as for the discovery cohort. To account for differences in sample quality and expected CNA resolution, normal cytology samples from healthy donors were processed at a coarser resolution, using a 500 kb bin size. This approach reduced noise and improved signal stability, which is appropriate given the lower DNA yield and complexity of normal samples. In contrast, samples from the HGSC group (tumors and cervical cytology samples) were analyzed at a finer resolution (50 kb bin size) to capture both focal and broad CNAs more accurately. The SRIQ-derived HCsig clusters were converted into centroids and used to perform a binary centroid-based prediction analysis (HGSC or healthy donors). Cross-tabulation was performed, showing the correct classification of 81/106 HGSC and 62/66 non-HGSC samples. Among women with HGSC and at least one cervical sample, 43/61 (70%) women had at least one HCpositive cervical sample (up to 98 months before diagnosis). In all, only four false positive and 25 false negative samples were found, yielding a specificity of 94% (95% CI: 0.90, 0.98) and sensitivity of 76% (95% CI: 0.70, 0.83). A receiver operating characteristic curve yielded an accuracy of AUC=0.83 (95% CI: 0.78, 0.89) (**Figure 5**). Notably, HCsig detected a majority of the early-stage HGSC cases (5/7) in the cervical samples from the external cohort.

**Figure 5.**
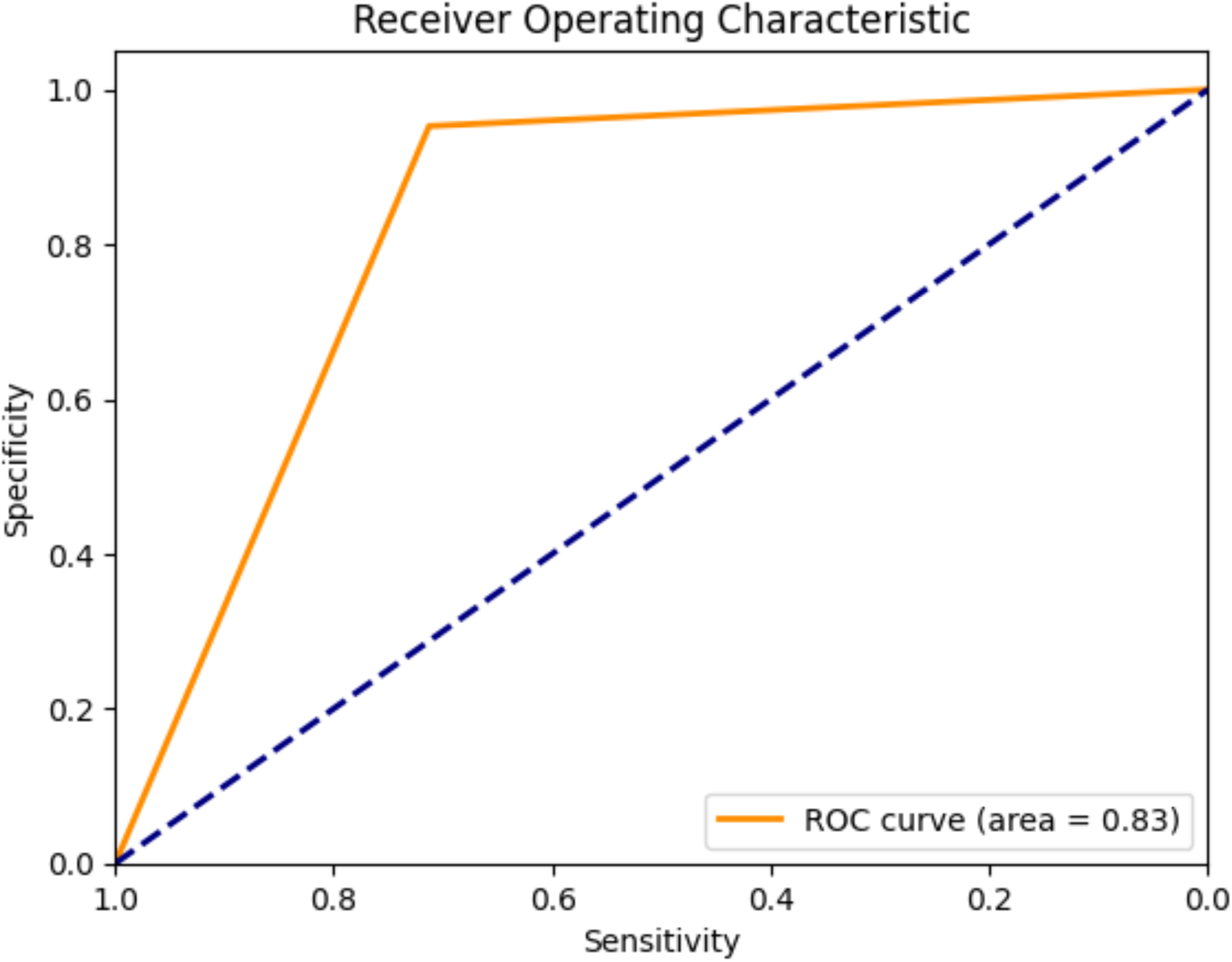
Validation of HCsig in the independent cohort. The receiver-operating characteristics curve (ROC) shows the sensitivity (x-axis) and specificity (y-axis) of HCsig in the independent cohort from Paracchini *et al.* (19) with healthy women and women with HGSC.

For benchmarking purposes, and to gauge the performance of HCsig in relation to an established metric, Fraction of Genome Altered (FGA) (21), we computed FGA values for all samples across both cohorts, showing inferior sensitivity, specificity and accuracy compared to HCsig (**Table S3**).

## DISCUSSION

In the present study we have demonstrated that the early detection of genomically aberrant HGSC-derived DNA in cervical samples collected from asymptomatic women is feasible. Increased early detection rates may enable earlier-stage diagnosis and hence improve survival rates for women with the most common type of ovarian cancer, HGSC. A screening tool for early detection of HGSC is an urgent unmet medical need and is required to be both highly sensitive and specific. Based on the premise that HGSC-derived cells and DNA can be shed into the cervix due to the anatomical connection via the uterine cavity and given evidence of genomic instability patterns in early HGSC development (7), we developed HCsig, a model leveraging copy number signatures from sWGS data for the early detection of HGSC in conventional cervical samples. While ultra-deep sequencing approaches have illustrated the possibility of highly sensitive detection of cancer-derived mutant *TP53* (11) (22), the occurrence of somatic clonal evolution linked to ageing limits its utility as a diagnostic test.

We show that sWGS combined with machine learning to characterize genomic features prototypical of HGSC and readily captured in cervical samples is both sensitive and specific, despite the low abundance of HGSC-derived DNA in the cervical samples. This approach provides a quantitative and qualitative measure of CNAs, presenting an improvement over previous purely quantitative measures like FGA (21) and gene-centric methods (8) (23) (24). Sampling in close proximity to the tumor mass contributes to the accuracy of this approach, supported by the higher fraction of true positive cervical samples vs. plasma samples from HGSC patients.

While retrospective in nature, the access to archival cervical samples allowed for an unbiased investigation of CNAs in non-symptomatic women up to several years before their HGSC diagnosis. The inability to correctly classify a subset of cervical samples from women with HGSC may be due to the closeness in time to surgery, involving multiple samplings during work-up, resulting in inferior sample quality or insufficient tumor-derived DNA. Another possible explanation may be blockage of the fallopian tubes in advanced-stage disease. Importantly, the performance of our model was validated in an independent cohort with high accuracy, suggesting that it is generalizable across populations and laboratories.

HGSC is characterized by a low frequency of recurrent oncogenic mutations, few recurrent CNAs and highly complex genomic profiles, which pose challenges for screening, diagnosis and classification of the disease. Novel approaches to early detection have focused on identifying the ubiquitous *TP53* mutations (9) (10) or a small panel of cancer-related genes (25) (24) in plasma or in samples of cervical or uterine origin. The main constraint of these mutation-based screening studies has been the lack of sensitivity, while specificity has recently emerged as the key challenge due to ultra-sensitive methods capturing clonal evolution of cancer-like age-related *TP53* mutations in healthy women and normal tissues (11). However, unlike mutations, CNAs are rarely present in normal tissues and may thus present a robust foundation for creating a test able to distinguish early cancer from benign or normal tissue. A quantitative analysis of genomic instability was recently proposed for early detection of ovarian cancer in cervical samples collected up to several years before diagnosis, with promising results (19). In the present study, we used an unsupervised approach, including mixture modelling and cosine similarities to extract both quantitative and qualitative genomic features known to occur in HGSC (15) for each cervical sample to generate a prediction model based on the distinct copy number features of HGSC. Importantly, the method is tumor mutation agnostic, able to capture early-stage disease, relatively inexpensive, and requires only small amounts of tumor-derived DNA. We provide proof-of-concept that CNAs found in archival cervical samples can be used as a biomarker to distinguish between HGSC and benign gynecological conditions, up to several years before the onset of symptoms and diagnosis. Of note, some women in the study underwent frequent sampling due to suspected cervical cell atypia, but these samples were generally called as HCnegative by the classifier, hence not confounding the results. Importantly, both advanced-stage and localized HGSC were detectable using our classifier, supporting the sensitivity and specificity of HCsig and the reported early genomic evolution of HGSC from STIC lesions (7). The detection of early-stage cancers, involving growth only on the ovaries and/or fallopian tubes, supports the robustness of the classifier and provides a promising approach for early detection with the possibility of increasing curability and reducing mortality. This supports the notion that sampling in close proximity to the tumor holds superior sensitivity over *e.g.* plasma and reflects the tubal origin of HGSC. Enrichment of copy number signatures indicative of mutational processes described in HGSC (15) and other epithelial cancers (16) (17), including HRD and tandem duplications support the HGSC-derived origin in the cervical samples in our study. In line with our findings, using a combination of whole-genome cell-free DNA (cfDNA) fragmentome and protein biomarker (CA-125 and HE4) analysis, Medina *et al.* recently reported a high accuracy for the detection of stage I-IV HGSC (26). Exploring our HCsig classifier in combination with fragmentomes in cervical samples and conventional protein biomarkers may improve the performance further.

A strength of our study was the inclusion of women with germline *BRCA1/2* mutations at high risk of developing HGSC, who underwent RRSO and hence a thorough pathology review. While no invasive cancers were detected, STIC lesions were noted for one of 13 women. Of note, 13/22 (59%) positive cervical samples from non-HGSC patients (‘false positives’) were from *BRCA1/2* mutation carriers, suggesting that *BRCA1/2* heterozygous cells may instigate carcinogenesis in the fallopian tubes due to potential haploinsufficiency. This possibility is supported by reports of *BRCA1* haploinsufficiency causing genomic instability in mammary epithelial cells (27, 28). There was a trend toward lower exposure to HCsig1 and HCsig3 in the HCsig positive samples from the RRSO group compared to the HGSC group with *BRCA1/2* mutations, potentially also supporting a haploinsufficient state in at-risk germline *BRCA1/2* mutant fallopian tube tissue. Reports suggest a prevalence of around 5-6% STICs in asymptomatic *BRCA1/2* mutation carriers (29) and the life-time risk of developing ovarian cancer for germline *BRCA1* and *BRCA2* mutation carriers is 44% and 17% without RRSO, respectively (30). An interval of several years from the occurrence of STIC lesions to the development of HGSC has been proposed (7, 29), indicating a potential window of opportunity for capturing loss of genomic integrity caused by heterozygous inactivation of *BRCA1* or *BRCA2* using HCsig. Furthermore, complex somatic CNAs can be found in early (stage I) ovarian cancers (31), corroborating HCsig as a tool for early diagnosis. Further exploration of the individual components of HCsig 1-5 in a larger cohort of asymptomatic women with germline *BRCA1*/*2* mutations until the time of RRSO in connection with detailed genomic and pathology review at the single cell level will allow for refinement of HCsig in the RRSO setting and to establish a cut-off for distinguishing between the *BRCA1/2* heterozygous state and complete loss of function. A recent report showed that *TP53* mutation burden in cervical cytology samples increased with age in *BRCA1/2* germline mutation carriers with HGSC (22), leading to greater genomic instability. This finding further supports the potential of risk assessment based on cumulative genomic instability using *e.g.* HCsig to aid in decision-making regarding the timing of RRSO for high-risk women.

## CONCLUSIONS

To summarize, we have shown that HGSC-derived copy number signatures can be captured with high sensitivity and specificity in cervical samples obtained from cervical cancer screening programs, including in samples collected up to several years before diagnosis, from women later diagnosed with HGSC, importantly including early-stage cancers. The results were validated in an independent, external cohort, strengthening the findings further and supporting the robustness and generalizability of the method. Our findings indicate that early-stage detection is possible in asymptomatic women, opening the possibility for the development of a copy number-based tool for early diagnosis. HCsig may be applied in a screening setting for early detection in high-risk individuals, and as a diagnostic tool in symptomatic women. Further characterization of the signatures and a large, prospective study should pinpoint which components to include in a diagnostic tool for early detection and increased survival.

## Data Availability

The data presented in this paper contain sensitive human information (whole genome sequencing) that cannot be shared openly. Processed data files and metadata necessary to reproduce the key findings are available at Mendeley under doi: 10.17632/d45hf6nxfv.2, in accordance with data-sharing and ethical guidelines.

https://data.mendeley.com/

## ABBREVIATIONS

ACE: Absolute Copy-number Estimation
ASCAT: Allele-Specific Copy Number Analysis of Tumors
*BRCA1/2*: Breast and Ovarian Cancer Susceptibility Protein 1/2
CA-125: Cancer Antigen 125 (Mucin 16)
cfDNA: cell-free DNA
CNA: Copy Number Alteration
FGA: Fraction of Genome Altered
FIGO: International Federation of Gynecology and Obstetrics
HCsig: High-grade serous ovarian cancer Cervical copy number signature
HE4: Human Epididymis Protein 4
HGSC: High-Grade Serous Cancer
HRD: Homologous Recombination Deficiency
RRSO: Risk-Reducing Salpingo-Oophorectomy
STIC: Serous Tubal Intraepithelial Carcinoma
sWGS: shallow Whole Genome Sequencing
*TP53*: Tumor Protein P53

## DECLARATIONS

### ETHICS APPROVAL AND CONSENT TO PARTICIPATE

The study was approved by the Ethics Committee at Lund University, Sweden (2014/717), with written informed consent for the prospective part of the discovery cohort and waiving the requirement for informed consent for the retrospective part of the discovery cohort.

### CONSENT FOR PUBLICATION

All authors approved the final manuscript.

### AVAILABILITY OF DATA AND MATERIALS

#### Code Availability

All analyses were performed using open-source software such as R, and Java programming language. The pipeline and other source codes used in this study can be found at: https://github.com/IngridHLab/HCsig, https://github.com/IngridHLab/CentriodClassification, https://github.com/sunnyveerla/SRIQ.

### AUTHORS’ CONTRIBUTIONS

Conceptualization: LMF, PK, IH

Resources: LMF, LP, LM, SM, MD, AM, SM, PK, IH

Data curation: LMF, SV, ML, AE, IH

Methodology: SV, ML, GT, AE, IH

Investigation: LMF, SV, AE, IH

Visualization: SV, AE

Funding acquisition: SM, MD, IH

Project administration: IH

Supervision: SV, IH

Writing – original draft: IH

Writing – review & editing: all authors

## ACKNOWLEDGMENTS

The authors wish to thank all patients who participated in the study, RCC South for providing infrastructure for the collection and biobanking of samples, SciLifeLab/Center for Translational Genomics (CTG) Lund University for sequencing services. We thank J-M Jönsson for critical review and feedback on the manuscript.

## AUTHORS’ INFORMATION

Laura Martin de la Fuente and Srinivas Veerla contributed equally to this work. Corresponding author: Ingrid Hedenfalk

## SUPPLEMENTARY INFORMATION

Supplementary Figures S1–S5 and Tables S1-S3

