## Supplementary Information for "Copy number signatures in cervical samples enable early detection of high-grade serous ovarian carcinoma"

Martin de la Fuente and Veerla, *et al.*

### Supplementary Figures

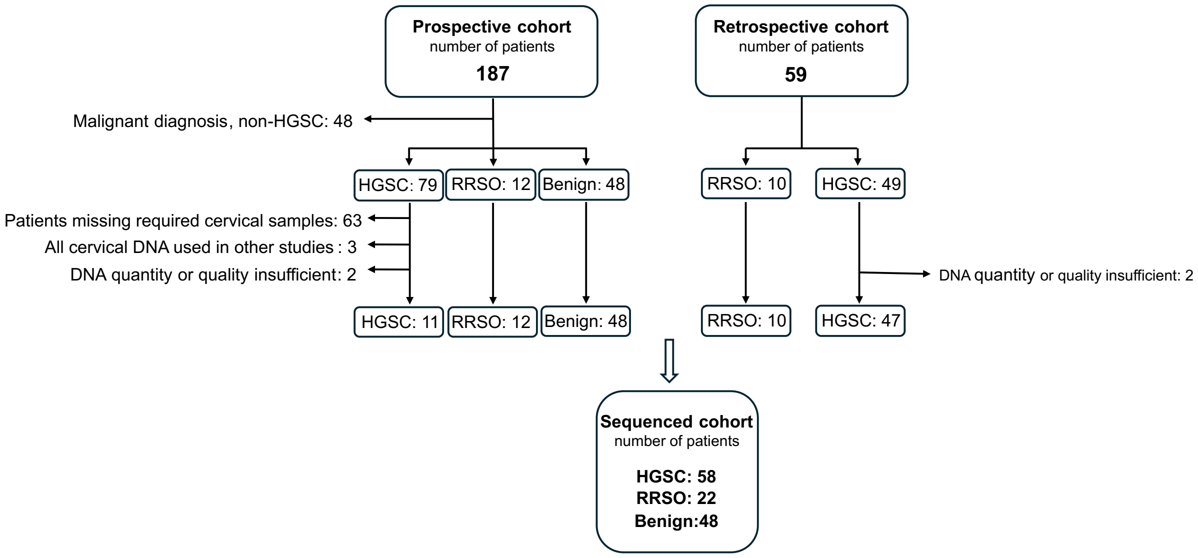

**Figure S1. Flowchart of patients included in the discovery cohort.** Patients from a previously reported prospective study [9] and a retrospective cohort were included in the study. Exclusion criteria are indicated.

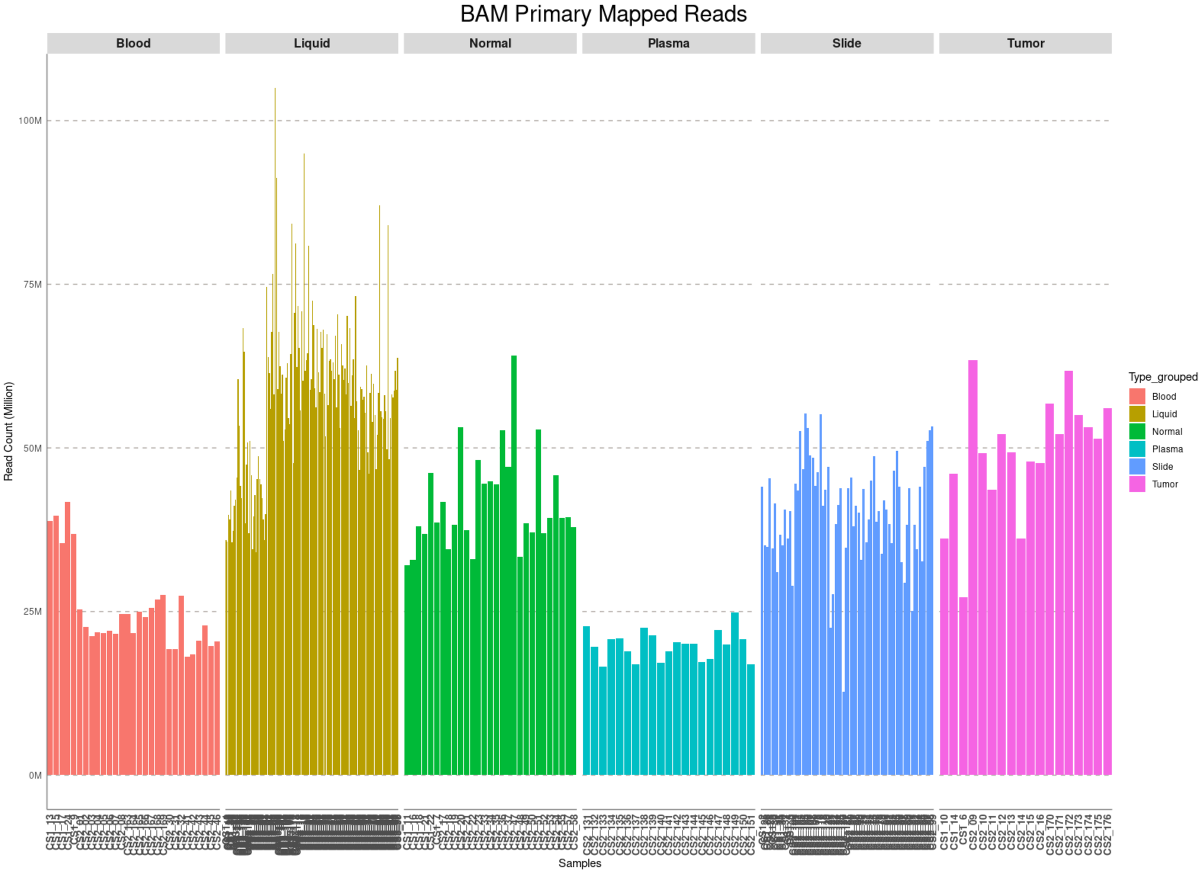

**Figure S2A:** Bar chart of average mapped read counts across all sample types.

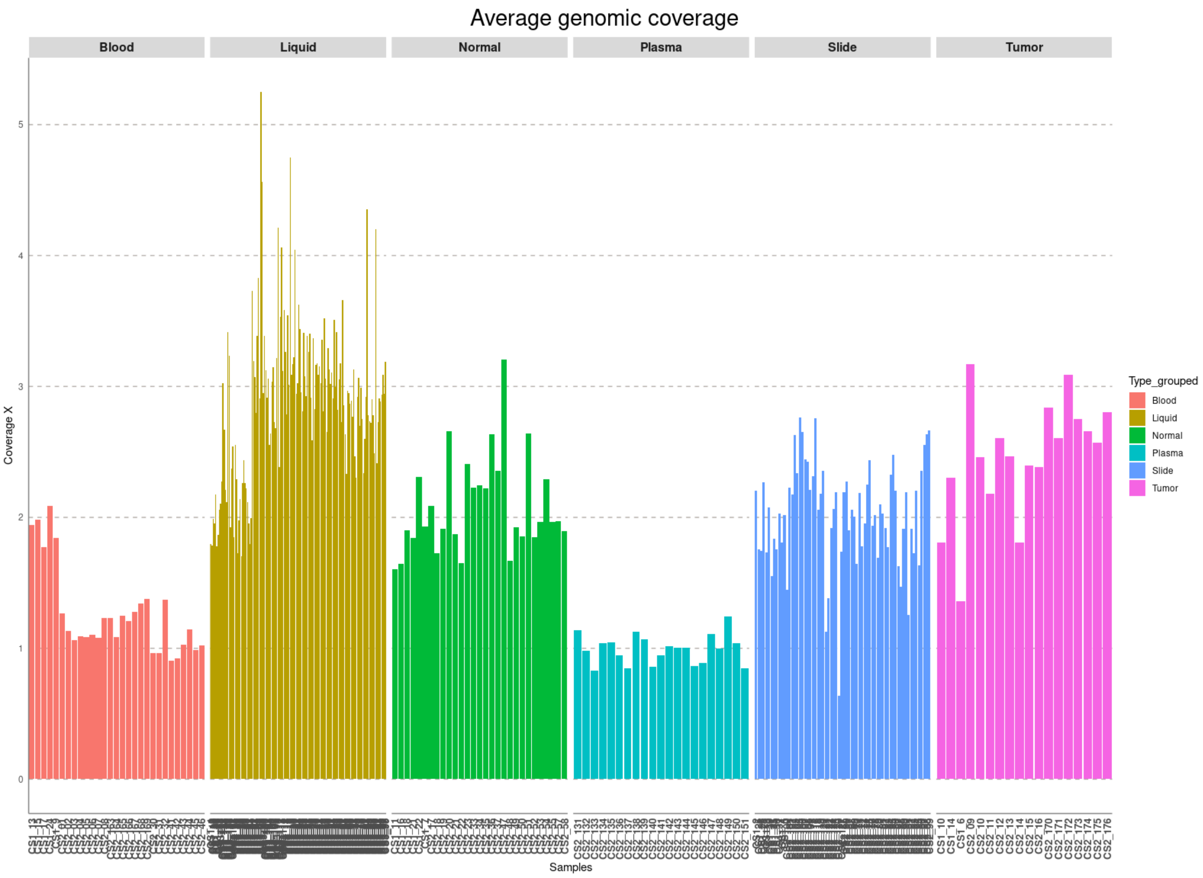

**Figure S2B:** Bar chart of average genomic coverage (X) across all sample types.

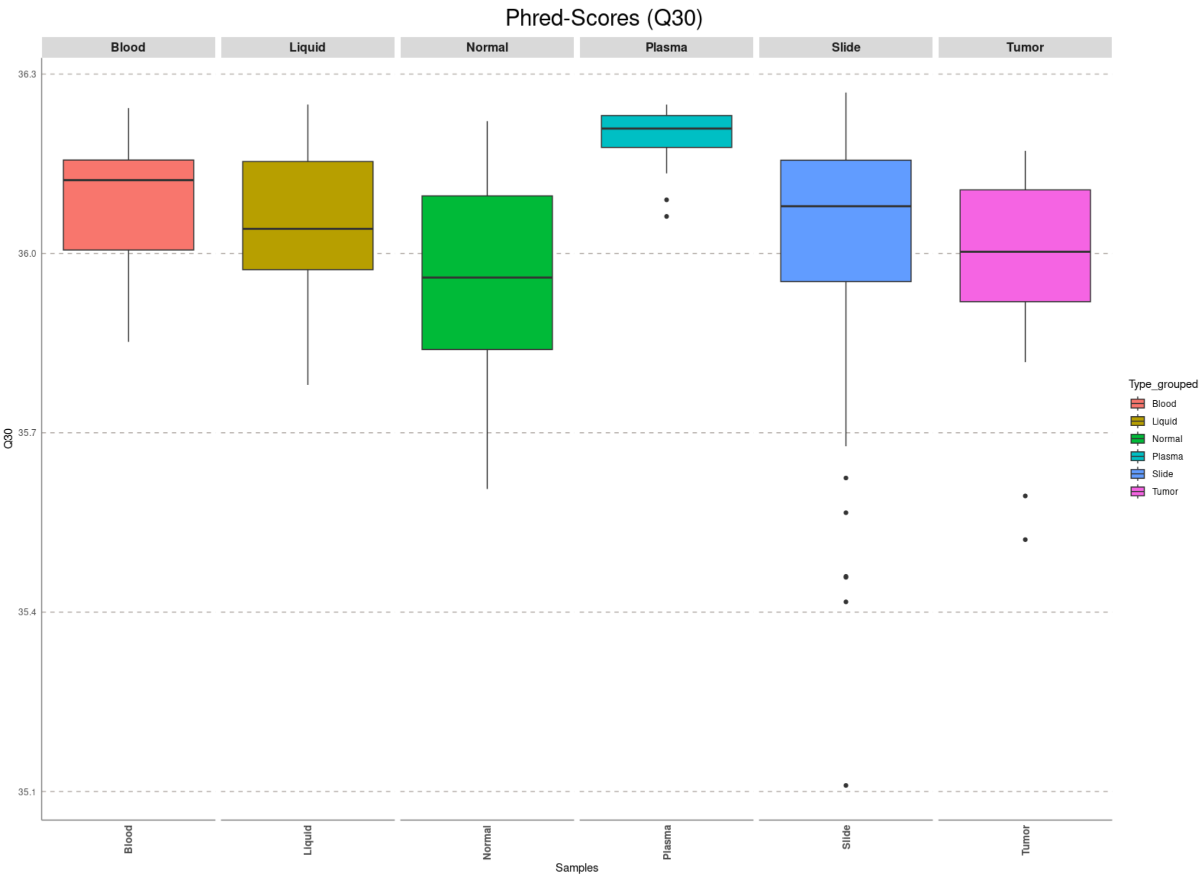

**Figure S2C:** Box plot of mean Phred scores (Q30) across all sample types.

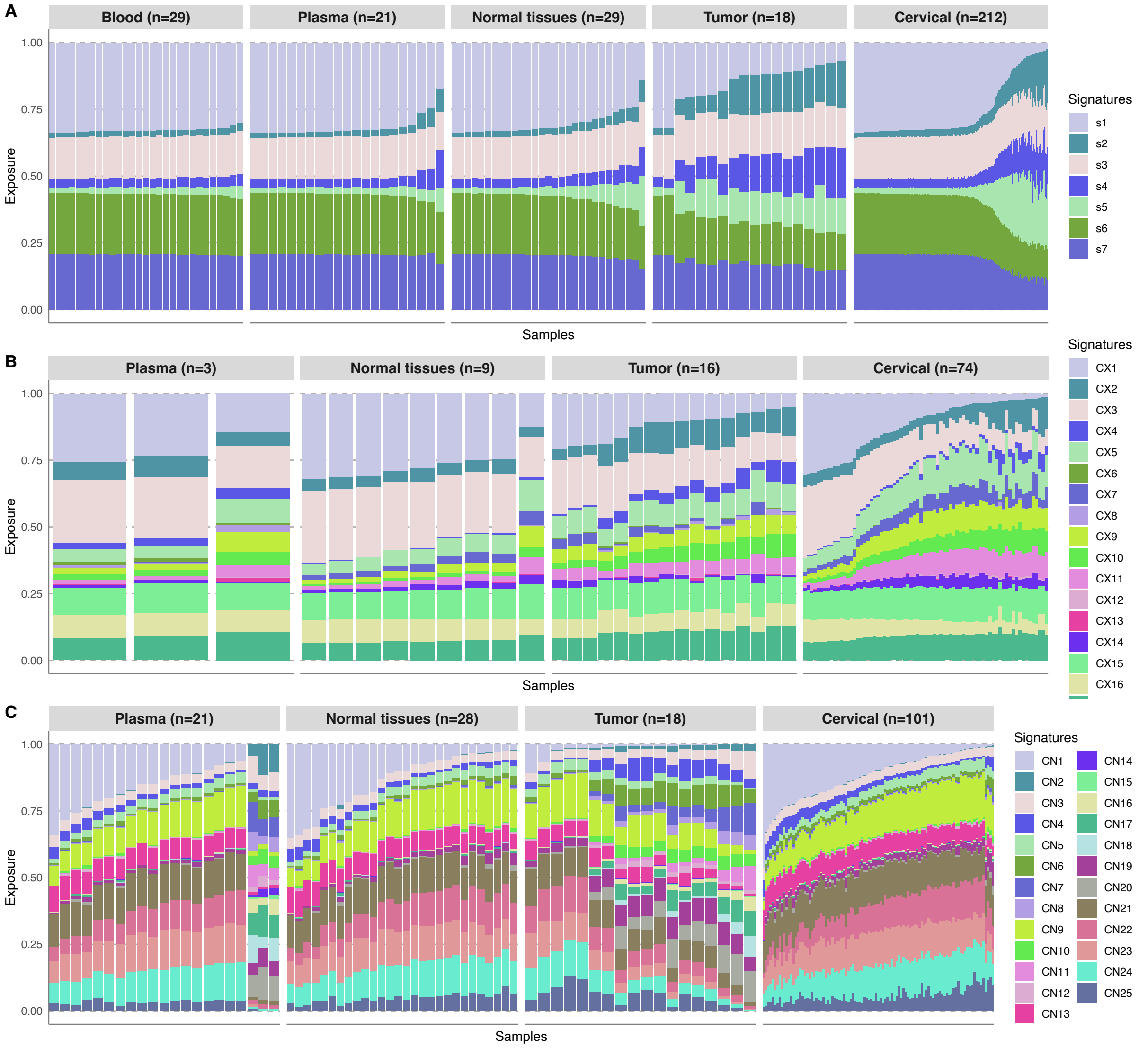

**Figure S3. Signature similarity exposure profiles for three cancer-derived copy number signatures across sample types. A.** Exposure profiles for HGSC-derived CN signatures from Macintyre *et al.* [15] for 309 samples. **B.** Exposure profiles for pan-cancer CIN signatures from Drews *et al.* [16] for 102 samples. **C.** Exposure profiles for panConusig signatures from Steele *et al.* [17] for 168 samples. Samples are ordered by type along the x-axes and cosine similarity values (0-1) are shown on the y-axes.

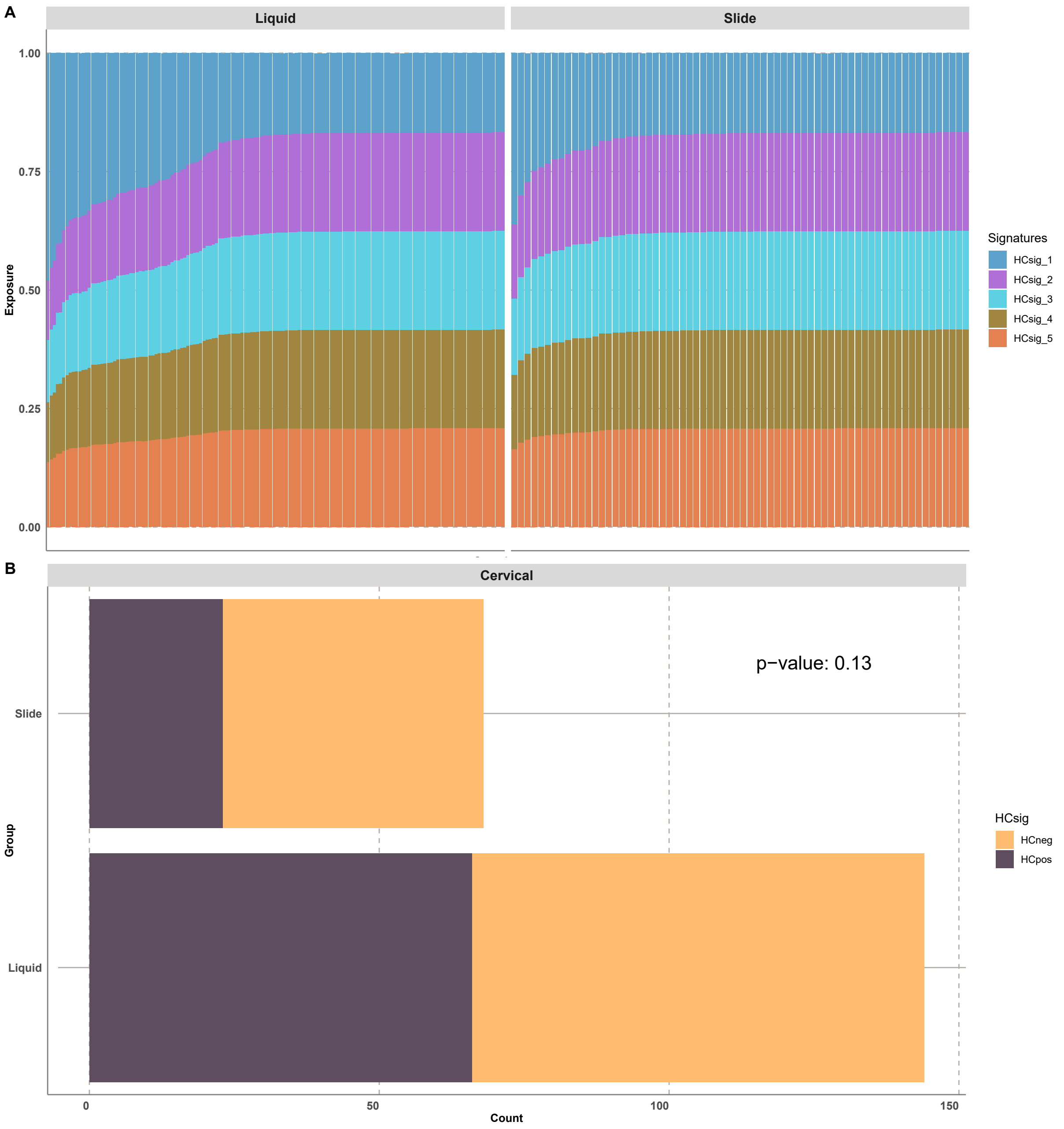

**Figure S4. HCsig signature exposure profiles for slide-based and liquid-based cervical samples. A.** Exposure profiles for HCsig1-5 for liquid and slide-based cervical samples. **B.** Classification of HCpositive and HCnegative for liquid and slide-based cervical samples.

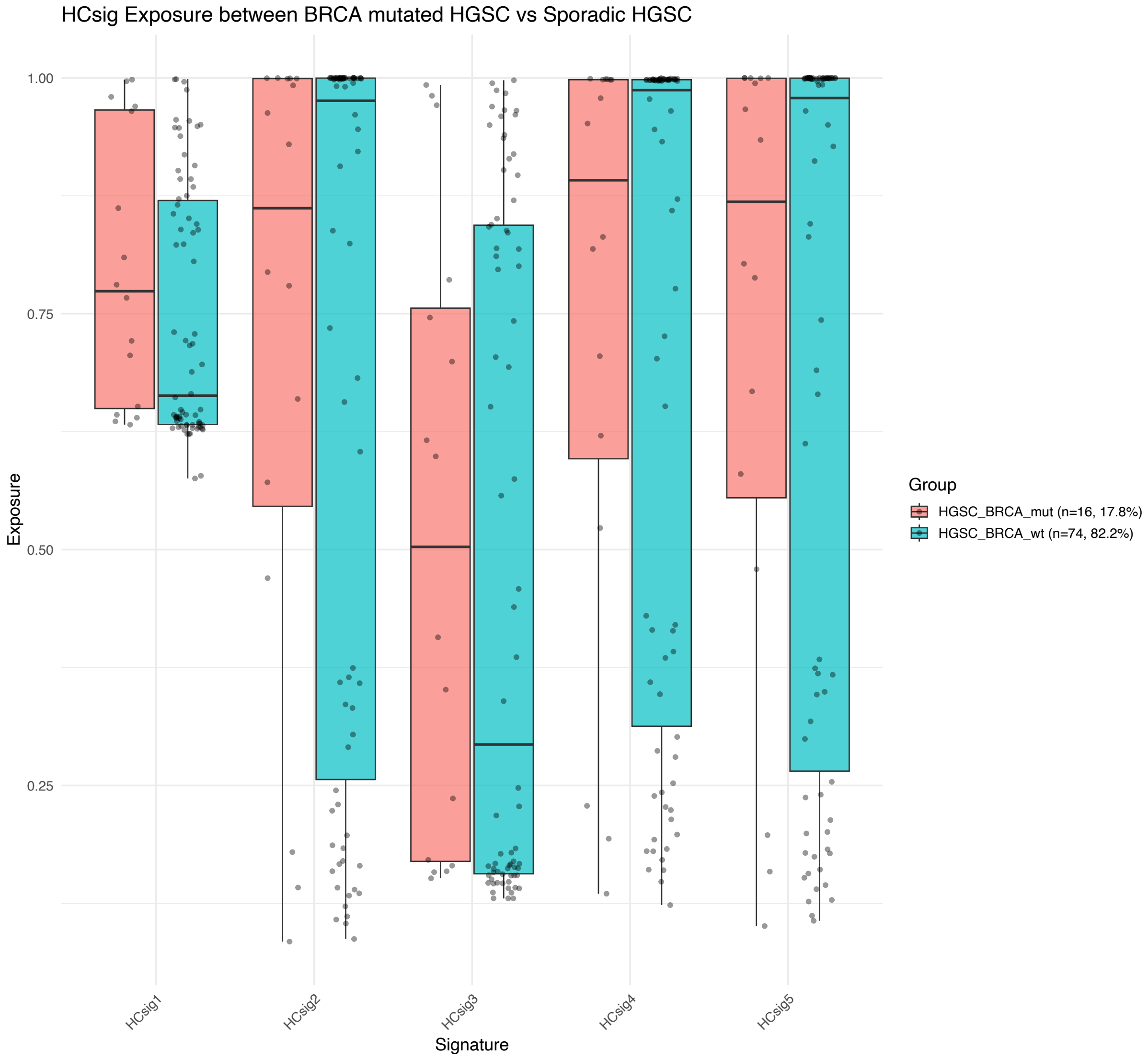

**Figure S5.** Box plots displaying signature exposures to HCsig1-5 in cervical samples from women with *BRCA1/2*-mutated HGSC and sporadic HGSC, respectively.

**Table S1. HCsig metadata.** Comprehensive information about patients, samples, and analysis results (separate file).

**Table S2. Overview of samples from the discovery cohort included in analyses of published cancer-derived copy number signatures.**

| | **Macintyre G** *et al* [15] | **Drews R** *et al* [16] | **Steele CD** *et al* [17] |
|---|---|---|---|
| **N = Samples (Patients)** | **309 (128)** | **102 (72)** | **168 (29)** |
| **HGSC** | **173 (58)** | **75 (47)** | **104 (18)** |
| Cervical Diagnostic | 31 (19) | 7 (6) | 21 (11) |
| Cervical Archival | 86 (52) | 48 (41) | 46 (17) |
| Tumor tissue | 18 (18) | 16 (16) | 18 (18) |
| Normal tissues | 9 (9) | 1 (1) | 8 (8) |
| Blood | 18 (18) | 0 (0) | 0 (0) |
| Plasma | 11 (11) | 3 (3) | 11 (11) |
| **RRSO** | **54 (22)** | **13 (12)** | **30 (4)** |
| Cervical Diagnostic | 17 (12) | 2 (2) | 8 (4) |
| Cervical Archival | 23 (14) | 8 (7) | 12 (4) |
| Tumor tissue | N/A | N/A | N/A |
| Normal tissues | 7 (4) | 3 (3) | 7 (4) |
| Blood | 4 (4) | 0 (0) | 0 (0) |
| Plasma | 3 (3) | 0 (0) | 3 (3) |
| **Benign** | **82 (48)** | **14 (13)** | **34 (7)** |
| Cervical Diagnostic | 55 (48) | 9 (8) | 14 (7) |
| Cervical Archival | 0 (0) | 0 (0) | 0 (0) |
| Tumor tissue | N/A | N/A | N/A |
| Normal tissues | 13 (7) | 5 (5) | 13 (7) |
| Blood | 7 (7) | 0 (0) | 0 (0) |
| Plasma | 7 (7) | 0 (0) | 7 (7) |

**Table S3. Fraction of Genome Altered (FGA).** Sensitivity, specificity and accuracy of FGA in the Discovery and Validation cohorts, respectively.

| | **Sensitivity**<br>% (CI) | **Specificity**<br>% (CI) | **Accuracy**<br>% (CI) |
|---|---|---|---|
| **Discovery cohort** | 69 (64-74) | 47 (42-53) | 49 (44-55) |
| **Validation cohort** | 62 (55-69) | 40 (33-47) | 59 (51-66) |
